# COVID-19 Implications of the Physical Interaction of Artificial Fog on Respiratory Aerosols

**DOI:** 10.1101/2021.03.18.21253891

**Authors:** Matthew Loss, Mark Katchen, Ilan Arvelo, Phil Arnold, Mona Shum

## Abstract

**Introduction:** Artificial fog is used in the film, television, and live entertainment industries to enhance lighting, as a visual effect, and to create a specific sense of mood or atmosphere. This study investigated whether the suspension time of respiratory aerosols spiked with tagged DNA tracers would change in the presence of glycerin- or glycol-containing artificial fogs.

**Methods & Materials:** Respiratory aerosols with tagged DNA tracers were sprayed into a closed environment without and with glycerin- or glycol-containing artificial fog, with air samples taken at regular intervals to determine the decay of tagged DNA tracer over time. The study treatments included Control (no fog), Glycerin Low (3 mg/m^3^), Glycerin High (∼15 mg/m^3^), Glycol Low (∼5 mg/m^3^), and Glycol High (∼40 mg/m^3^).

**Results:** All artificial fog treatments had lower mean log reduction curves compared to the Control treatment. Compared to the Control and Glycerin Low treatments, the differences in mean log reduction for nearly all other artificial fog treatments were statistically significant (p<0.001); the difference between Control and Glycerin Low treatments was not statistically significant (p=0.087). The differences in mean log reduction between treatments using the same artificial fog type were not statistically significant.

**Conclusion:** Artificial fog use does not increase suspension time of respiratory aerosols, and therefore does not appear to increase the risk of airborne transmission of diseases from respiratory aerosols, such as COVID-19. Of the two types of artificial fogs investigated, that containing glycol decreased suspension time more than that containing glycerin. In practice, the additional reduction in suspension time provided by the physical interaction of respiratory aerosols with artificial fog does not suggest any practical benefit for using artificial fog as a control measure.

## 1 Introduction

Artificial fog is used most often for creating special effects in the film, television, and live entertainment industries to make lighting or lighting effects visible, and to create a specific sense of mood or atmosphere. Devices, referred to as fog machines, work by either condensing vapor generated by heating liquid fogging fluid, or by mechanically generating aerosols directly from liquids. The fog consists of small liquid aerosols suspended in air. The aerosols include the same ingredients as the fluids used in the machines, which are water-based, but often combined with a percentage of glycerin, glycols, or highly-refined mineral oils. The fog is not real smoke, soot, or char. It is not generated by thermal decomposition or burning of fluid ingredients, although a small amount of thermal decomposition byproducts may be produced during the process of heating the fluid prior to condensation.

With the onset of the severe acute respiratory syndrome coronavirus 2 (SARS-CoV-2) pandemic, now termed as Coronavirus Disease 2019 (COVID-19), there has been a concern expressed within these entertainment industries regarding the interaction of artificial fog and respiratory aerosols, which may contain and transmit COVID-19. The aerosol transmission of COVID-19, in the absence of artificial fog, in well-ventilated indoor spaces is not an efficient route of transmission for the virus based on modeling of the COVID-19 aerosol (Smith et al., 2020), as COVID-19 microdroplets, owing to their small size, contain less virus than the larger droplets, known as respiratory aerosols. Respiratory aerosols, generated by coughing, sneezing, or speaking, tend to fall to the ground within approximately three feet (one meter) of the generating source. The question arises whether the physical interaction of artificial fog on respiratory aerosols could increase suspension of these larger aerosols containing more virus and thus increase the likelihood of COVID-19 transmission and subsequent infection. Specifically, this current study investigated whether the suspension time of respiratory aerosols spiked with tagged DNA tracers would change in the presence of glycerin- or glycol-containing artificial fogs.

## 2 Methods and Materials

### 2.1 DNA Tracers

The tagged DNA tracers, supplied by SafeTraces Inc., were housed in and sprayed by Flairosol spray bottles. The DNA tracer solutions were approximately 1% solids to mimic saliva. The tagged DNA tracers used by SafeTraces are generally recognized as safe (GRAS) by qualified experts when aerosolized in this type of application, and when aerosolized, they are well below the U.S. Occupational Safety and Health Administration’s exposure limit for particulates not otherwise regulated. The Flairosol spray bottle produced a median particle size (D50) of 87.27 micrometers (µm) (+/- 1.62 µm) with a distribution ranging from 43.25 µm on the 10^th^ percentile to 191.36 µm on the 90^th^ percentile; the volume mean diameter (if all particles were the same sized spheres) was on average 103.87 µm (+/- 1.92 µm). Therefore, the spray bottle reproduced respirable aerosols and droplets that are similar in size and distribution compared to those generated by sneezing, coughing, and talking (Xie et al., 2007; Xie et al., 2009). Due to these factors, and the low detection limit achievable, SafeTraces Inc. and their tagged DNA tracers were deemed safe and adequate in simulating respiratory aerosols.

### 2.2 Study Design Summary

Respiratory aerosols with tagged DNA tracers were sprayed into a closed environment with and without artificial fog, where air samples of aerosols were taken at regular intervals to determine the decay of tagged DNA tracer over time. A small office boardroom measuring 545 cubic feet (8’11” long by 8’ 4” wide by 7’ 5” high), occupied with one table and two chairs, was sealed along the walls, door, window, supply air diffuser, and ceiling with one millimeter-thick poly sheeting (HDX, Canada). This poly sheeting created a closed environment where airflow in or out of the room was minimized, thereby limiting tagged DNA tracer decay due to natural settling processes only. Five treatments were completed: one control treatment, two glycerin-containing artificial fog treatments, and two glycol-containing artificial fog treatments. The two glycerin-containing artificial fog treatments aimed to maintain airborne glycerin concentrations at approximately 1.5 milligrams per cubic meter (mg/m^3^) or 15 mg/m^3^; the two glycol-containing artificial fog treatments aimed to maintain airborne glycol concentrations at approximately 5 mg/m^3^ or 40 mg/m^3^. These glycerin and glycol concentrations aligned with regulatory or guideline limits commonly used for workplaces in North America (i.e., for 12-hour time-weighted average and ceiling limits, respectively). For each treatment, six trials were completed; each trial consisted of spraying a unique tagged DNA tracer into the room and collecting one five-minute sample every five minutes from the time of spray until thirty-minutes had elapsed, for a total of six samples collected per trial and 36 samples per treatment.

### 2.3 Air Sampling

Two Pilot Studies were conducted to refine the method to ensure proper set-up of equipment and capture of the decay of tagged DNA tracer (Figure 1). Each sample consisted of a Grade A-E 25-millimeter (mm) glass fiber filter (Sterlitech Corporation, USA) housed in a 50 mm long, three-piece conductive black polypropylene cassette housing cowl with a backing pad (Zefon International, USA) attached to a Leland Legacy Pump via Tygon^®^ tubing (Saint-Gobain Performance Plastics Corp., USA). The Leland Legacy Pump was pre-calibrated to draw air at approximately 8 liters per minute using a Defender 510 (Mesa Labs, USA) with the first sample. The cowl was angled downward at approximately 45 degrees and suspended approximately five feet above the ground in the middle of the room by an aluminum tripod (Environmental Monitoring Systems, USA). The cassette angle helped minimize collection of aerosols through deposition and mimicked the human nose more accurately when used in conjunction with the cowl. Once the first sample was ready, the tagged DNA tracer fluid was sprayed five times from a Flairosol spray bottle, distributing the aerosols into each corner and center of the room; the different directions of each spray assisted in homogenizing the aerosol in the room quickly. Immediately after spraying, the Leland Legacy Pump was turned on to begin the first sample; the first sample started after the sprays owing to Researcher limitations. Once this first sample started, a table fan with a blade diameter of twelve inches (GD Midea Environment Appliance Mfg. Co., Ltd, China) located in the Southeast corner was turned on to its lowest speed (660 feet per minute (ft/min) at the face, 275 ft/min at a distance five feet away) and oscillated over a 90 degree range from the Southwest to Northeast corners; operation of the fan began after the sprays to ensure it did not disrupt the initial natural dispersion of aerosols but helped homogenize the aerosols in the room afterwards. After a sample duration of five minutes, the Leland Legacy Pump was paused, sampled cassette was removed, a new cassette was attached to the Tygon^®^ tubing, and then the Leland Legacy Pump was restarted; it took approximately ten seconds to complete sample swapping. The same Leland Legacy Pump was used to ensure the flow rates and pump parameters were consistent between each sample. This process was repeated for each subsequent sampling time (interval): 5 to 10 minutes, 10 to 15 minutes, 15 to 20 minutes, 20 to 25 minutes, and 25 to 30 minutes. After all six samples were completed for a given trial, the last sample was used to post-calibrate the Leland Legacy Pump.

**Figure 1.**
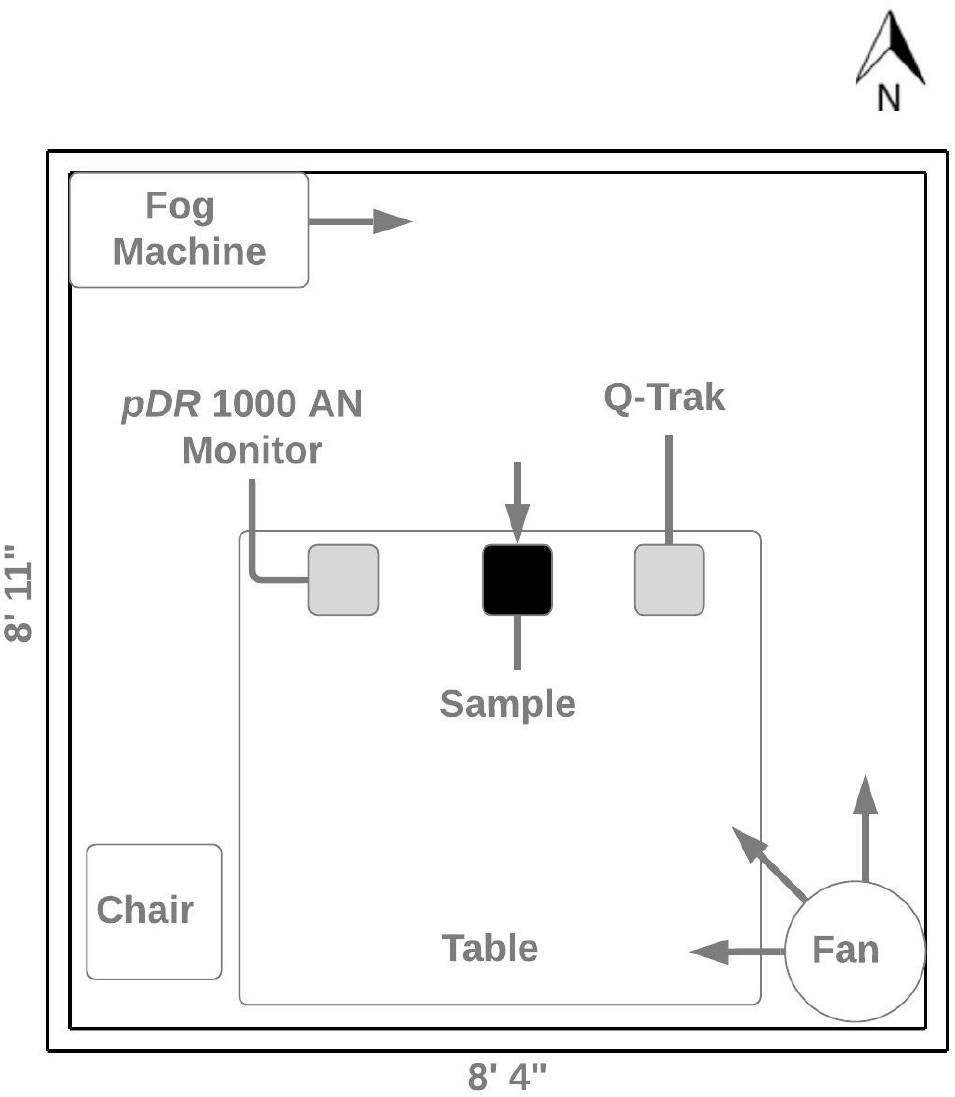
Study design physical layout for performing air sampling. The arrows indicate the direction of air movement away from the fan and fog machine, and the direction of air movement when collecting air samples.

### 2.4 Artificial Fog

Water-Vapor Haze™ (CITC, USA) was used in a Haze Max machine (CITC, USA) to generate the glycerin-containing artificial fog treatments. SmartFog™ Fogging Fluid: 3 Minute Low-Ground Fog (CITC, USA) was used in a Fog Max machine (CITC, USA) to generate the glycol-containing artificial fog treatments. The fog machines were turned on and dispensed fog until the desired airborne glycerin or glycol concentration was reached. A personal DataRAM™ *p*DR-1000AN Monitor (Thermo Fisher Scientific Inc., USA) placed next to the samples was adjusted using a calibration factor of 1.87 (Environ International Corporation, 2014) to measure glycerin aerosols and 0.66 (Environ International Corporation, 2002) to measure glycol aerosols; calibration factors adjusted the instrument’s sensors to specifically measure glycerin or glycol aerosols. This instrument was moved around the room periodically to ensure homogeneous glycerin and glycol concentrations. Before each treatment, the instrument was zero calibrated and programmed to record every ten second average concentration throughout the treatment. The *p*DR-1000AN Monitor has an aerodynamic particle cut point range at 10 µm and a concentration measurement range from 0.001 to 400 mg/m^3^. The Researcher inside the room encouraged dispersion of the artificial fog by manually fanning the air with a clipboard. When fanning, care was taken to not fan air upwards towards the sample being collected. Once the desired concentration was reached, the tagged DNA tracer and sample collection process started. Periodically throughout the sampling period, the fog machine dispensed artificial fog in 0.5 to 1.5 second bursts, followed by dispersion via fanning, to maintain a consistent glycerin or glycol concentration in the air. The same process and actions were repeated with the Control treatment, except distilled water was used in the fog machine instead of a glycerin- or glycol-containing artificial fog.

### 2.5 Temperature and Relative Humidity

Temperature in Celsius (°C) and relative humidity in percentage (%) were measured continuously during every sample using a Q-Trak Model 7565 with Probe 982 (TSI, USA), with every ten second average reading recorded. The instrument probe was located next to the samples in the middle of the room.

### 2.6 Sampling Shipment

All trials for a treatment were completed in the same day. A unique tagged DNA tracer was used for each trial to eliminate possible cross-contamination between trials. At the end of each treatment, each sampled filter was removed from its cowl using clean plastic tweezers and placed into a 2 milliliter (mL) DNA LoBind Tube (Eppendorf AG, Germany), then placed into a 2-millimeter thick plastic bag. All samples were shipped to SafeTraces Inc. (Pleasanton, California, USA) for laboratory analysis. Bulk liquid samples of each tagged DNA tracer used were collected by pouring 2 mL of the fluid into a 2 mL DNA LoBind Tube and placing into a 2-millimeter thick plastic bag. The floor, walls, ceiling, table, and chairs of the closed environment and plastic tweezers were cleaned with a 10% bleach solution at the end of each treatment.

### 2.7 Quality Control

An OmniAire 1200PAC Portable Air Cleaner (Omnitech Design, USA) was operated overnight for approximately sixteen hours at medium speed to filter the air between treatments to minimize cross-contamination between different treatments, because the same set of tagged DNA tracers were used for each treatment. Approximately three field blanks per treatment were collected for quality assurance and quality control purposes to evaluate sample handling and potential routes of contamination. Each field blank was treated the same as samples, except no air was drawn through them.

### 2.8 Laboratory Analysis

Filter samples contained inside 2 mL DNA LoBind Eppendorf Tubes were stored at SafeTraces Inc. in a -20°C freezer prior to starting the DNA extraction protocol. Samples were then taken out of the freezer and allowed to equilibrate for approximately 10 minutes to room temperature (21°C). A volume of 0.5 mL of elution buffer was added into the 2 mL tube containing the filter samples, vortexed at full speed for 2 minutes using a VortexGennie2, then centrifuged using a minifuge at 10,000 revolutions per minute (rpm) for 10 seconds to pool the eluate at the bottom of the tube. A 4 microliter (µL) sample of the eluate was transferred to the corresponding reaction well of a 0.2 fast 96-well non-skirted PCR plate which contained 16 µL of master mix reagents (IDT prime time gene expression master mix, water, primers, and SYBR green) per well. The 96-well was sealed using a MicroAmp Optical Adhesive Film, centrifuged using an Eppendorf centrifuge 5810 at 4,000 rpm for 1 minute. The qPCR plate containing a 20 µL total reaction volume per well (4 µL sample with 16 µL master mix) was then loaded into a QuantStudio3 or QuantStudio5 qPCR instrument operated following these thermal cycling parameters: activation step of 95°C for 1 minute, then 40 cycles of 95°C for 0.1 second and 60°C for 20 seconds of annealing time using the standard FAM 2-step fast qPCR protocol. Readings were collected at the end of the annealing/extension step. The QuantStudio platform generated a quantification cycle (Cq) value associated with the input DNA concentration. The Cq value was then used to estimate the number of DNA copies in the reaction well using a standard curve.

### 2.9 DNA Tracer Quantification

The number of DNA copies aerosolized were calculated and adjusted based on the concentration of DNA measured in each bulk liquid sample (Equation 1). This value was also adjusted based on an aerosol fraction, which synchronizes the Flairosol spray bottle aerosol fraction to that generated from sneezing, talking, and coughing. Given the described distribution of aerosol sizes generated from sneezing, talking, and coughing, and the potential for partial or total evaporation of liquid aerosols between 60 to 100 µm (Xie et al, 2007), an aerosol diameter cut-off point of73.56 µm was selected. This cut-point corresponded to 37.35% volume of the distribution of aerosols released by the Flairosol spray bottle.

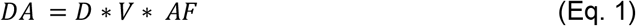

DA = number of DNA copies aerosolized

D = number of DNA copies per milliliter

V = sprayed volume in milliliters

AF = aerosol fraction, assumed to be 37.35 percent (0.3735)

The logarithmic (log) reduction was calculated for each sample using base ten (Equation 2). In addition to log reduction, the number of copies per million copies aerosolized was calculated for each sample (Equation 3).

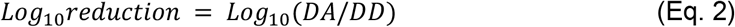

DA = number of DNA copies aerosolized

DD = number of DNA copies detected

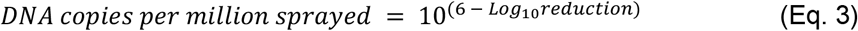

### 2.10 Mean Log Reductions

For each treatment, the mean log reduction, standard deviation, sample size, and 95% confidence interval were calculated for each sampling time. This analysis was repeated for the number of DNA copies per million sprayed. For each treatment, the mean log reduction and 95% confidence interval were plotted against the sampling time, with the x-axis for sampling time and y-axis on a log scale for log reduction and number of DNA copies per million sprayed, yielding a mean log reduction curve for each treatment.

### 2.11 Temperature and Relative Humidity Analysis

The mean temperature, relative humidity, and artificial fog concentration were calculated for each sample, sampling time, trial, and treatment. The mean differences in these variables were calculated and compared between all treatments, between all sampling times, and between trials within each treatment.

### 2.12 Statistical Analysis

All data were organized using Microsoft Excel (Microsoft Corporation, 2018); statistical analyses and figures for log reductions were conducted and produced in R version 4.0.3 (R Core Team, 2021) using packages contained in Tidyverse (Wickham et al., 2019).

The assumption of normality for the treatments was qualitatively assessed, because the sample size was too small for formal statistical tests. The assumption of homogeneity of variance was tested using a Bartlett Test of Homogeneity of Variance (Bartlett, 1937), applied to the combined levels of the variables “Treatment - Interval”. A two-way analysis of variance (ANOVA) (Chambers & Hastie, 1992) was performed with the levels of the variables “Treatment” and “Interval” to determine if there was any significant interaction between the two variables. An ANOVA and Tukey Honest Significant Differences test (Miller, 1981; Yandell, 1997) was performed for “Treatment” and “Interval” to determine if mean differences in overall log reductions were statistically significant. For all statistical analyses, a significance level of 5% was used to reject the null hypothesis (αα = 0.05).

## 3 Results

### 3.1 Summary

Sampling was completed between November 2020 and January 2021 in Burnaby, British Columbia, Canada. All treatments were completed by the same Researcher, in the same office space, under similar environmental conditions. The maximum mean difference in temperature and relative humidity between treatments was 3.6°C and 12.4%. Two trials were excluded from the analysis: one was a calibration trial to refine the methodology (Glycerin Low, Trial 1) and the other was analyzed for the incorrectly tagged DNA tracer (Glycol Low, Trial 3). Nearly all treatments with artificial fog maintained glycerin or glycol concentrations near the desired concentration. One exception is the Glycerin Low treatment, where the glycerin concentration was higher (Table 1).

**Table 1.**
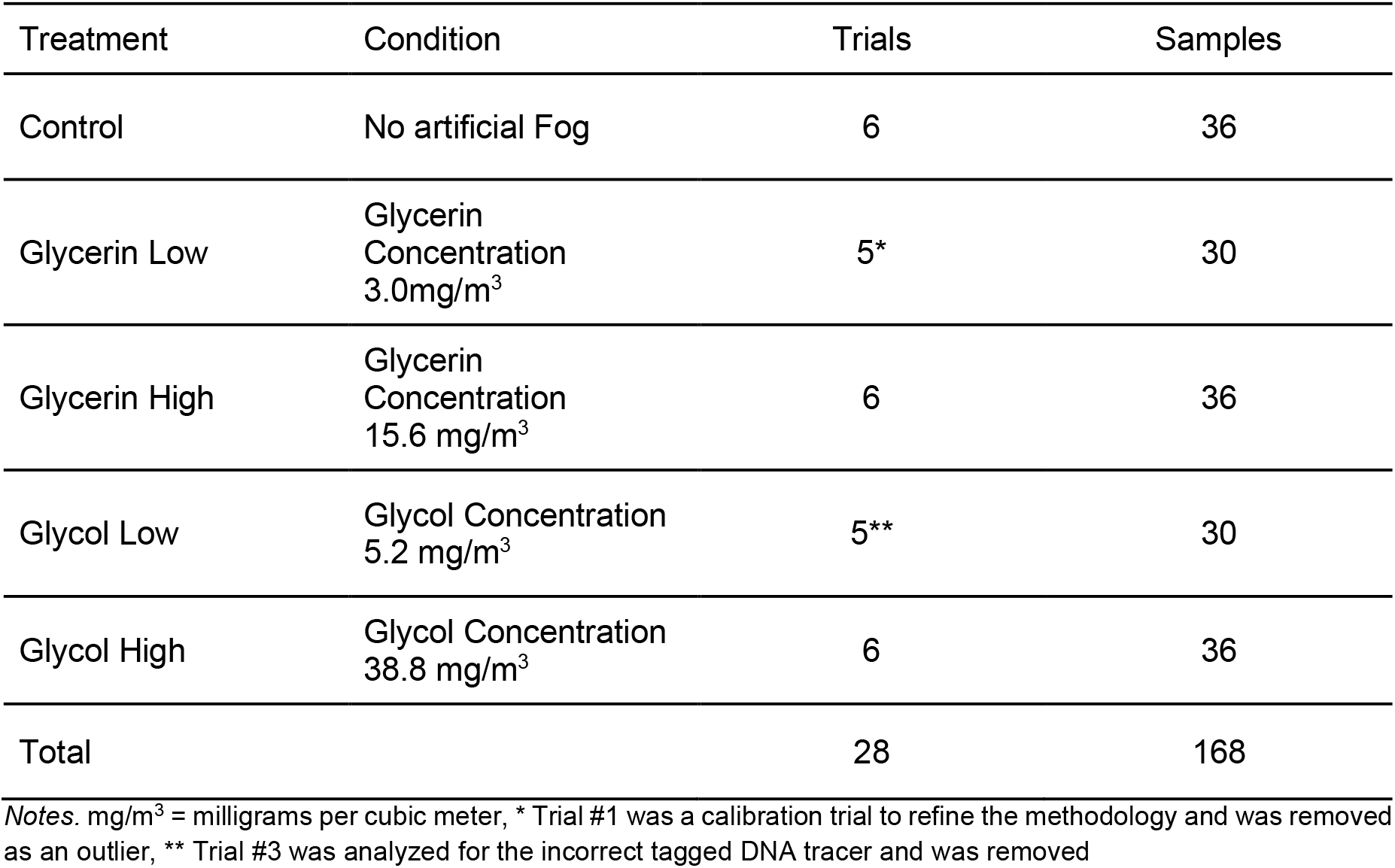
Summary of Sampling Completed.

| Treatment | Condition | Trials | Samples |
| --- | --- | --- | --- |
| Control | No artificial Fog | 6 | 36 |
| Glycerin Low | Glycerin Concentration<br>3.0mg/m<sup>3</sup> | 5* | 30 |
| Glycerin High | Glycerin Concentration<br>15.6 mg/m<sup>3</sup> | 6 | 36 |
| Glycol Low | Glycol Concentration<br>5.2 mg/m<sup>3</sup> | 5** | 30 |
| Glycol High | Glycol Concentration<br>38.8 mg/m<sup>3</sup> | 6 | 36 |
| Total |  | 28 | 168 |

Notes. mg/m<sup>3</sup> = milligrams per cubic meter, \* Trial #1 was a calibration trial to refine the methodology and was removed as an outlier, \*\* Trial #3 was analyzed for the incorrect tagged DNA tracer and was removed

### 3.2 Suspension Time

All artificial fog treatments had lower mean log reduction curves compared to the Control treatment, indicating the tagged DNA tracers in air decayed at a faster rate, and their suspension time in air was shorter (Figure 2). The Glycol High mean log reduction curve was the lowest, with the shortest suspension time of tagged DNA tracers in air. The glycol-containing fog treatments had lower mean log reduction curves compared to the glycerin-containing fog treatments. The Glycerin High mean log reduction curve was lower than the Glycerin Low mean log reduction curve.

**Figure 2.**
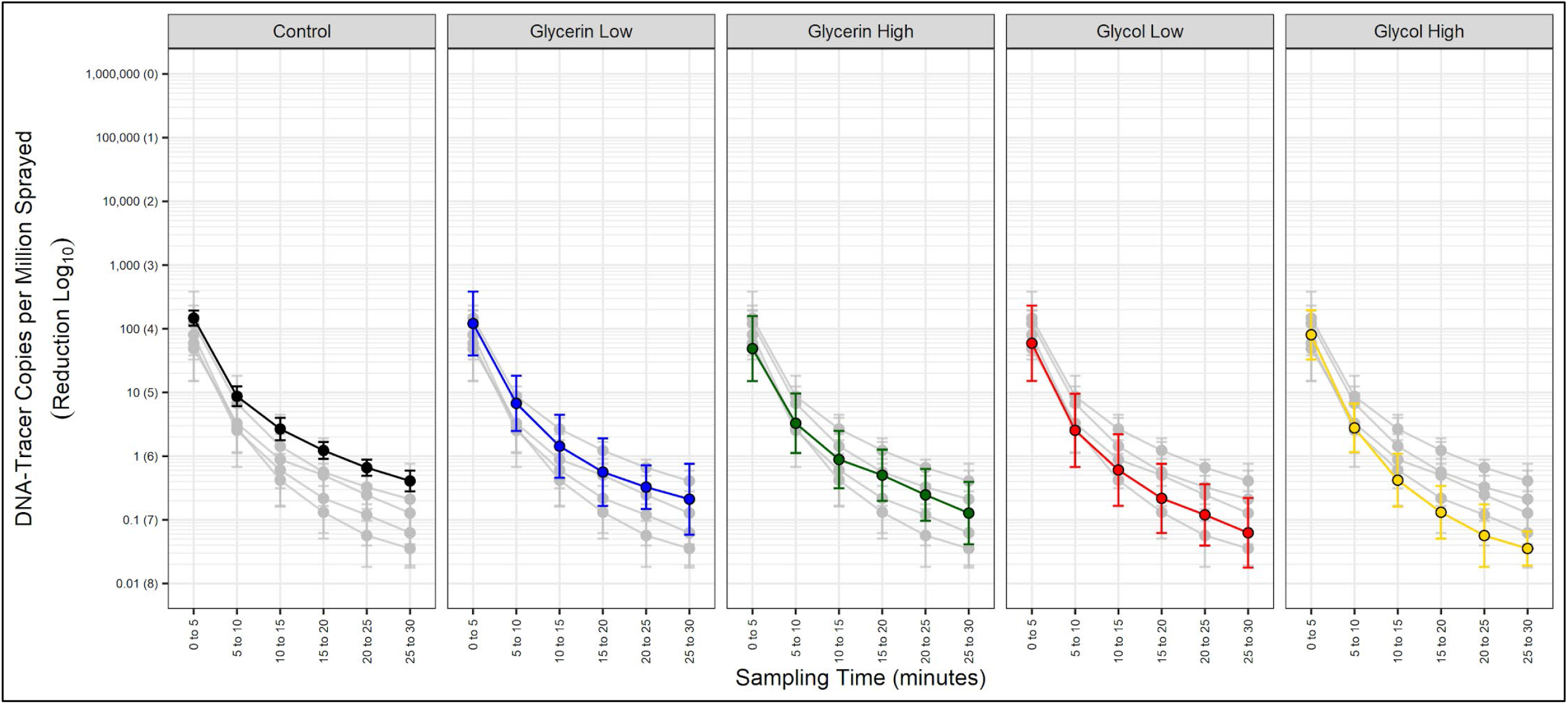
Mean log reduction of tagged DNA tracers in air over time with and without artificial fog. For each treatment, the mean log reduction (mean ± 95% CI) was calculated and plotted at each sampling time (interval). The solid point represents the mean and the bar and whiskers represent the 95% confidence interval around the mean. Within a treatment panel, the colored line represents that treatment’s mean log reduction curve while the grey curves represent all other treatment mean log reduction curves.

The overall mean log reduction, from the time of spray until 30 minutes had elapsed, ranged from 6.4 logs for the Control treatment to 7.5 logs for the Glycol High treatment. Within the first and last measured sampling times (intervals), the total log reduction measured for the Control treatment was 2.6 logs. The artificial fog treatments resulted in reductions ranging from 2.8 to 3.4 logs, with Glycol High yielding the largest overall log reduction. In general, with each successive sampling interval, the magnitude of reduction decreased for all treatments. The largest mean log reductions for all treatments occurred during the first three sampling times, which were the first 15 minutes after spraying. Between 15 to 30 minutes, the total mean log reduction was 1.1 logs or less for all treatments.

Given the sample size, no formal statistical test was applied to test the assumption of normality for the log reductions. Based on a qualitative assessment of the individual data points, the data follows a central trend; therefore, this assumption is moderately accurate. The test for homogeneity of variance applied to the combined levels of the variables “Treatment-Interval” yielded no statistically significant differences (p = 0.11, K-squared = 38.37). The two-way ANOVA determined the interaction between the variables “Treatment” and “Interval” was not statistically significant (p = 0.633), indicating that there is no interaction between the variables, and their effects on mean log reduction are independent of each other. When analyzed independently, the effect of “Treatment” was statistically significant (p<0.001), and the effect of “Interval” was also statistically significant (p<0.001).

Compared to the Control and Glycerin Low treatments, the differences in mean log reduction for nearly all other artificial fog treatments were statistically significant (p<0.001) (Table 2). The difference in mean log reduction between Control and Glycerin Low treatments was not statistically significant (p = 0.087). The differences in mean log reduction between treatments using the same artificial fog type were not statistically significant.

**Table 2.** Tukey Honest Significant Difference Comparing Treatment Mean Log Reductions.

| Treatment Comparison | Mean Difference<br>in Log Reduction | 95% CI of the Mean<br>Difference |  | p-value |
| --- | --- | --- | --- | --- |
|  |  | Low | High |  |
| Glycerin Low vs Control | 0.23 | -0.02 | 0.49 | 0.087 |
| Glycerin High vs Control* | 0.45 | 0.21 | 0.69 | <0.001 |
| Glycol Low vs Control* | 0.65 | 0.39 | 0.90 | <0.001 |
| Glycol High vs Control* | 0.78 | 0.53 | 1.02 | <0.001 |
| Glycerin High vs Glycerin Low | 0.22 | -0.04 | 0.47 | 0.129 |
| Glycol Low vs Glycerin Low* | 0.41 | 0.15 | 0.68 | <0.001 |
| Glycol High vs Glycerin Low* | 0.54 | 0.29 | 0.80 | <0.001 |
| Glycol Low vs Glycerin High | 0.20 | -0.06 | 0.45 | 0.209 |
| Glycol High vs Glycerin High* | 0.33 | 0.08 | 0.57 | 0.003 |
|  |  | Low | High |  |
| Glycol High vs Glycol Low | 0.13 | -0.12 | 0.38 | 0.617 |

Notes. \* statistically significant, CI = confidence interval

The differences in mean log reduction between nearly each sampling time were statistically significant (p<0.05); the exception is between sampling times “20 to 25” and “25 to 30”, where the difference in mean log reduction was not statistically significant (p = 0.18).

## 4 Discussion

It was shown that artificial fog appears to decrease suspension time to varying degrees depending on the chemical composition and airborne concentration. The largest decrease in suspension time was observed after the first five minutes, where all treatments had at least a four-log reduction. The log reduction observed in the artificial fog treatments was, in general, statistically significant compared to the Control treatment. The magnitude of reduction past four logs became exponentially smaller with each additional log reduction. A change from four to five logs is equivalent to a reduction of an additional 0.009%, and a change from five to six logs is equivalent to a reduction of an additional 0.0009%. Although artificial fog treatments were observed to have lower mean reduction curves, the amount of additional reduction yielded from the physical interaction of artificial fogs does not appear to be practically significant as a control measure for reducing airborne aerosols.

The natural decay of artificial fog in air was semi-quantitatively assessed by bringing up fog levels to both high and low levels and observing the decay with the *p*DR-1000AN Monitor. To decay 90% from a starting concentration that matched the “High” treatment, it took the glycerin-containing artificial fog approximately one hour and seventeen minutes and the glycol-containing artificial fog approximately ten minutes. The presence of the tagged DNA tracer in air did not appear to drastically alter this decay duration. The intended use of these two artificial fogs were different, where the glycerin-containing artificial fog was designed to stay suspended in air to create a haze effect, while the glycol-containing artificial fog was designed to be a low-lying fog. The different purposes of each artificial fog may have contributed to their effect on respiratory aerosol suspension time. If the artificial fog physically interacts with respiratory aerosols, artificial fogs that are designed to fall more quickly out of air, or be low-lying, may physically remove respiratory aerosols and shorten suspension time compared to artificial fogs designed to remain in air longer. The opposite does not appear to be true, as the artificial fogs designed to stay in air did not increase respiratory aerosol suspension time in air compared to no artificial fog.

The relative humidity during the Glycol High treatment had a mean difference of -12.4% compared to the Control treatment, noticeably lower than the other artificial fog treatments. Lower relative humidity promotes increased desiccation of aerosols in air. One previous study identified that with decreasing relative humidity, the total mass of aerosols with a mean aerodynamic diameter of 2.5 micrometer in air increases, meaning the suspension time increases (Zhaou et al., 2020). Given the mean differences in temperature and relative humidity between the Glycol High treatment (22.0°C and 64.1%) and Control treatment (21.7°C and 76.6%), the aerosol suspension time during the Glycerin High treatment is estimated to increase by approximately less than 1% based on the work performed by Zhaou et al. (2020). Additionally, Chen and Zhao (2010) determined that the influence of temperature and relative humidity on the dispersion of droplets with an initial diameter range of 0.1 to 200 µm was negligible. It is possible the differences in temperature and relative humidity may have affected the Glycol High treatment mean log reduction curve, but the impact is not expected to meaningfully alter its relationship with the Control treatment mean log reduction curve. The same is true for the other artificial fog treatments.

The limitations of this study are noteworthy. The small sample size for each treatment was limited, which impacted the resolution of mean log reduction curves and reduced the power to detect statistically significant differences in mean log reductions. Despite this limitation, there was consistency within each treatment and sampling interval, with all mean standard deviations being less than 0.50. The samplers used were not size selective, thereby may have captured all aerosol size fractions and potentially captured larger aerosols outside the respiratory size range. This limitation was partially controlled for by adjusting the calculated number of DNA copies to align the Flairosol spray bottle aerosol distribution with the distribution of aerosols generated by sneezing, talking, and coughing and which partially or totally evaporated. This study did not investigate how artificial fog may affect the propagation distance of respirable aerosols, nor the disinfection properties of glycerin or glycol on tagged DNA tracers. Only one type of each glycerin-containing and glycol-containing artificial fog fluid was used for this study. There are a large range of manufacturers and fluid types available, each with slightly different liquid compositions and percentages of glycerin or glycol. The impact of different liquid compositions and percentages of glycerin or glycol were outside the scope of this study.

## 5 Conclusion

This study supports that artificial fog does not increase the suspension time of respiratory aerosols in air, but rather has no effect or decreases the suspension time. Of the two types of artificial fogs investigated, artificial fog containing glycol decreased suspension time more than that containing glycerin. Regardless of the type of artificial fog used, suspension time decreased more with increasing artificial fog concentration, albeit not statistically significantly. In practice, the additional reduction in suspension time provided by the physical interaction of respiratory aerosols with artificial fog does not suggest any practical benefit for using artificial fog as a control measure. The principal outcome supported by this study was that artificial fog use does not increase suspension time of respiratory aerosols, and therefore does not appear to increase the risk of airborne transmission of diseases from respiratory aerosols, such as COVID-19.

## Data Availability

Data may be available upon request, and is provided on a case by cases basis.

## Acknowledgements

Thank you to the International Alliance for Theatrical Stage Employees (IATSE), Local 891, Directors Guild of Canada – BC (DGC-BC), and the Alliance of Canadian Cinema, Television and Radio Artists’ Union of British Columbia Performs (UBCP/ACTRA) for their financial support of this study. Thank you to SafeTraces Inc. for the tagged DNA tracers, sampling equipment, and laboratory analysis support. Thank you to CITC and Omnitec Design for supplying the artificial fog fluids, fog machines, and OmniAire 1200PAC Portable Air Cleaner.

